# APOE4 carriers display loss of anticipatory cerebral vascular regulation over AD progression

**DOI:** 10.1101/2024.10.11.24315344

**Authors:** Jacqueline A. Palmer, Carolyn S. Kaufman, Alicen A. Whitaker-Hilbig, Sandra A. Billinger

## Abstract

**INTRODUCTION:** Maintenance of cerebral blood flow during orthostasis is impaired with aging and associated with cognitive decline, but the effect of Apolipoprotein L4-allele (*APOE4*) is unknown.

**METHODS:** Older adults (n=108) (*APOE4* carriers, n=47; noncarriers, n=61) diagnosed as cognitively-normal (NC), MCI, or AD participated. Middle cerebral artery blood velocity (MCAv), assessed using Transcranial Doppler ultrasound, and beat-to-beat mean arterial blood pressure (MAP) were continuously recorded during a sit-to-stand transition. Anticipatory and orthostatic-induced MCAv and MAP responses were compared between genotypes and across disease progression.

**RESULTS:** Cognitively-normal *APOE4* carriers showed greater anticipatory MCAv increase, greater MCAv decrease with orthostasis, and shorter latency of peripheral MAP responses to orthostasis compared to noncarriers. MCAv and MAP responses were delayed and attenuated across the *APOE4* disease progression, with no differences between genotypes in MCI and AD.

**DISCUSSION:** *APOE4* carriers and noncarriers present with distinct phenotypes of cerebral vascular dysfunction during hemodynamic orthostatic challenge. Unique cerebral and peripheral vascular compensation observed in *APOE4* carriers may be lost as AD progresses.

## 1. Introduction

Possession of the *Apolipoprotein* L4-allele (*APOE4*) is an established central player in the pathogenesis of Alzheimer’s disease.[1–4] Increasing evidence points to impairments in cerebral vascular function and a greater decline in cerebral blood flow (CBF) in *APOE4* carriers that may contribute to cognitive impairment and dementia.[5–9] Cognitively-normal older adults who carry the *APOE4* allele demonstrate a greater decline in CBF with aging compared to noncarriers,[10] and an earlier blood-brain barrier breakdown that predicts subsequent cognitive decline.[9] Cognitively-normal older adult *APOE4* carriers with lower cerebral vascular function and greater amyloid-beta deposition have lower cognitive executive function, an interaction that is not present in noncarriers.[11,12] Interestingly, differences in cerebral vascular function have even been detected in young adult *APOE4* carriers, who show reduced cerebrovascular reactivity to carbon dioxide compared to noncarriers.[13] Together, these results implicate differences in cerebral vascular function between *APOE* genotypes that may influence their vulnerability to Alzheimer’s disease.

Regulation of cerebral blood flow (CBF) velocity under states of hemodynamic challenge, such as those induced during orthostasis, can be measured using transcranial Doppler ultrasound (TCD)[14–16]. Cerebral blood flow responses during orthostasis offer insight into function of the cerebral vascular system not gleaned from resting states alone, particularly in older adult populations [17] and diseased brain states.[18] In response to hypotension induced during transition from seated to standing positions, the cerebral microvasculature typically dilates quickly to increase blood flow to the brain.[19] Impaired ability to regulate the cerebral pressure/flow relationship results in repeated, exaggerated drops in cerebral perfusion that could damage neuronal tissue over time.[20,21] Abnormalities in cerebral blood flow responses to orthostasis have been observed in older adults[17] and a range of diseased brain states (e.g. stroke,[22,23] concussion,[24,25] and diabetes[26]); however, relationships to cognitive impairment and dementia have been inconsistent, with some studies finding no differences in orthostatic-induced cerebral blood flow decreases in MCI and AD and others show subtle differences.[27–30] One study found that transgenic *APOE4*-expressing mice have reduced cerebral blood flow and an inability to increase cerebral blood flow to meet the demands of active brain areas; this resulted in local hypoxia causing white matter damage and cognitive dysfunction.[8] These findings suggest that *APOE4* may impair vasodilatory mechanisms involved in cerebrovascular regulation, which are necessary for effective responses during orthostasis, potentially leading to downstream damage to the brain parenchyma.

In addition to differences in cerebral vascular health and function, emerging evidence suggests that *APOE4* carriers may display earlier signs of neurovascular compensation during cognitive and motor tasks compared to their noncarrier (*APOE3 or APOE2*) peers.[8,31] For example, cognitively-normal older adults who carry *APOE4* show greater cognitive-motor dual-task interference during gait,[31] and animal models of *APOE4* show an impaired ability to match cerebral blood flow with increases in task-related brain activity compared to *APOE3*.[8] Greater prefrontal cortical activity, implicated in cognitive dual-task interference, can be engaged in an anticipatory manner prior to movement initiation.[32] In addition to cognitive processing, the prefrontal cortex has also been strongly implicated in the regulation of cardiovascular function.[33,34] However, whether or how differences in cortically-mediated whole-body behaviors may interact or influence differences in cerebral vascular function observed between *APOE4* carriers and noncarriers is unclear.

Considering differences in neurovascular brain function between *APOE4* carriers and noncarriers are detected even at a young age, yet only half of heterozygous *APOE4* carriers develop AD,[35] there must be contributing neuroprotective factors that influence Alzheimer’s disease development. Here, we hypothesized that older adult *APOE4* carriers would show a dysfunctional cerebrovascular response to orthostasis compared to their noncarrier peers, but would also display vascular compensation during the preclinical stages of AD progression. We further hypothesized that disease progression to MCI and AD would be characterized by greater cerebrovascular dysfunction and loss of vascular compensation in *APOE4* carriers. We tested the effect of *APOE4* genotype on anticipatory and orthostatic-induced changes in cerebral blood velocity and beat-to-beat peripheral mean arterial blood pressure (MAP) in a group of older adults classified as cognitively-normal (NC), mild cognitive impairment (MCI), or early-stage Alzheimer’s disease (AD).

## 2. Materials and Methods

### 2.1. Participants

Participants (n=108) were diagnostically classified as cognitively-normal (NC, n=65), mild cognitive impairment (MCI, n=25), or early Alzheimer’s disease (AD, n=18) (**Table 1**). Inclusion criteria for the present analyses were (1) age 65-90 years, (2) absence of neurologic diagnosis other than MCI or AD, (3) ability to follow two-step verbal commands, (4) presence of a TCD signal, and (5) absence of orthopedic disability to prevent independent standing. Exclusion criteria were (1) insulin-dependent diabetes, (2) peripheral neuropathy, (3) active coronary artery disease and congestive heart failure. The experimental protocol was approved by the University of Kansas Institutional Review Board (IRB#: STUDY 00147888 and 0011132) and all participants provided written informed consent.

### 2.2. Clinical neuropsychological test battery

All participants completed a standard in-person clinical and cognitive evaluation on a separate day, during which the Clinical Dementia Rating (CDR) scale and the United States Alzheimer’s Disease Research Center network neuropsychological test battery was performed by a trained clinician and psychometrist, respectively.[36,37] Clinical and cognitive data were reviewed and each participant was classified as being cognitively-normal (NC), having mild cognitive impairment (MCI), or Alzheimer’s disease (AD) at a consensus diagnostic conference[38]. Participants also completed a Mini-Mental State Exam (MMSE)[39] and Montreal Cognitive Assessment (MoCA)[40](**Table 1)**.

### 2.3. Sit-to-stand protocol and data acquisition

TCD was used to assess middle cerebral artery blood velocity (MCAv) during a sit-to-stand positional transfer. A 2-MHz TCD probe (RobotoC2MD, Multigon Industries) was used to record right MCAv over the temporal window. The left MCA was used if the right MCA signal was absent. Continuous beat-to-beat MAP was recorded through a cuff around the left middle finger (Finapres Medical Systems, Amsterdam, The Netherlands). A 5-lead electrocardiogram (Cardiocard; Nasiff Associates, Central Square, New York) continuously recorded heart rhythm and was used to synchronize MCAv and MAP across the cardiac cycle.[41,42] A capnograph (BCI Capnocheck Sleep 9004; Smiths Medical, Dublin,OH) recorded continuous expired end tidal carbon dioxide (P_ET_CO_2_) through a nasal canula and participants were instructed to breathe through their nose throughout the 3-minute duration of the sit-to-stand recording. All data were recorded at 500Hz. During the first minute of the recording, the participant remained seated quietly. At the 60-second mark of the recording, the experimenter verbally cued the participant to stand and remain standing for 2 minutes. Time-synchronized raw data were acquired through an analog-to-digital unit (NI-USB-6212, National Instruments) and custom written MATLAB software (The Mathworks Inc. Natick, MA).

### 2.3. Quantification of anticipatory and autonomic responses

Recordings of MCAv and MAP were visually inspected and discarded when R-to-R intervals were >5 Hz or changes in peak MCAv or MAP exceeded 10 cm/s or 10mmHg in a single cardiac cycle, respectively. Trials with <85% samples were discarded from analysis. Mean MCAv and MAP were calculated from the area under the curve (AUC) for each cardiac cycle.[43] The onset of the sit-to-stand event was identified at 60 seconds into the recording, and the onset beat=0 was identified as the beat immediately following t=60s. Two mean baseline (BL) metrics were computed within the 30 beats immediately preceding the onset of the sit to stand transition, in which BL1= −31 to −16 beats and BL2=−15 to −1 beats, and onset =beat 0. Automated identification of the post-stand MCAv and MAP nadir (lowest point after standing) and latency in seconds from the onset time=0 to nadir were identified within the first 20 beats immediately following the onset of sit-to-stand, and were visually confirmed for accuracy. We calculated the % change in anticipatory ((BL2-BL1)/BL1*100%)) and orthostatic post-stand responses ((nadir-BL2)/BL2*100%)).

### 2.4. *APOE* genotyping

Taqman single nucleotide polymorphism (SNP) allelic discrimination assays (ThermoFisher) were used to determine *APOE4, APOE3*, and *APOE2* alleles to the two *APOE*-defining SNPs, rs429358 (C_3084793_20) and rs7412 (C_904973_10) using whole blood samples stored at −80 degrees Celsius.[44,45] Individuals were classified as an *APOE4* carrier in the presence of 1 or 2 *APOE4* alleles (e.g. E3/E4, E4/E4). Individuals with homozygous E3 (i.e. E3/E3) or E2/E3 were classified as noncarriers.

### 2.5. Statistical analyses

We tested for normality and heterogeneity of variance of all data used for analyses using Kolmogorov-Smirnov and Levene’s tests, respectively. First, we compared acute MCAv and MAP change during orthostasis between cognitively-normal *APOE4* carriers and noncarriers. We used separate two-way repeated measures analysis of variance (RM-ANOVAs) with factors of genotype (*APOE4* carrier, noncarrier) and time (BL1, BL2, post-stand) for each MCAv and MAP. We compared MCAv and MAP anticipatory and orthostatic response amplitudes, as well as orthostatic response latencies between cognitively-normal *APOE4* carriers and noncarriers using independent t-tests and within time using paired t-tests.

We then used two-way ANOVAs to compare acute MCAv and MAP change during orthostasis over the course of Alzheimer’s disease progression in each *APOE4* carriers and noncarriers. We performed RM-ANOVA tests with factors of diagnosis (NC, MCI, AD) and time (BL1, BL2, post-stand) for each genotype separately. Two-way independent ANOVAs were used to compare the magnitude of MCAv and MAP anticipatory responses, orthostatic responses, and orthostatic response latencies within and between *APOE4* carriers and noncarriers. All analyses were performed using SPSS version 29 with an a priori level of significance set to 0.05.

## 3. RESULTS

For two participants (NC, *APOE4*, n=1; AD, *APOE4*, n=1), MAP data were unavailable due to technical issues with the device and were discarded from this part of the analysis. Two different participants (NC, *APOE4*, n=1; AD, *APOE4*, n=1), had <85% samples available free of artifact on TCD signals and were discarded from this part of the analysis.

Within each diagnostic group (NC, MCI, AD), there were no significant differences between *APOE4* carriers and noncarriers in age (p>0.58), MMSE (p>0.236), or MOCA score(p>0.693). In the NC group, there was a greater proportion of females in the noncarrier compared to the *APOE4* carrier genotype (p=0.030), with no sex differences in MCI or AD diagnostic groups (p>0.065). We observed no main effect of time (BL1, BL2, or post-stand) in P_ET_CO_2_ (*p*=0.988) and no differences in P_ET_CO_2_ or heart rate between *APOE4* carriers and noncarriers or diagnosis group at any time point (*p*>0.408).

### 3.1. Effect of *APOE* genotype on cerebral and peripheral vascular responses to orthostasis

Cognitively-normal *APOE4* carriers showed a greater anticipatory increase and greater orthostatic-induced decrease in MCAv compared to noncarriers (**Figure 1A**). There was a significant time-by-genotype interaction (F_2,124_ = 4.43, p=0.014); while there were no between-group differences in absolute MCAv at any time point (p>0.285), *APOE4* carriers showed a significant within-group anticipatory increase in MCAv between BL1 and BL2 (p=0.004) that did not occur in noncarriers (p=0.125) (**Figure 1A**). Both groups showed a significant decrease in MCAv between BL2 and post-stand (p<0.001) (**Figure 1A**). When comparing the normalized magnitude of change in MCAv between groups, *APOE4* carriers demonstrated a greater anticipatory increase (p=0.002) (**Figure 1C)** and greater post-stand reduction in MCAv compared to noncarriers (p=0.023) (**Figure 1D**). No group differences in MCAv nadir latency were observed (p=0.147) (**Figure 1E**).

**Figure 1.**
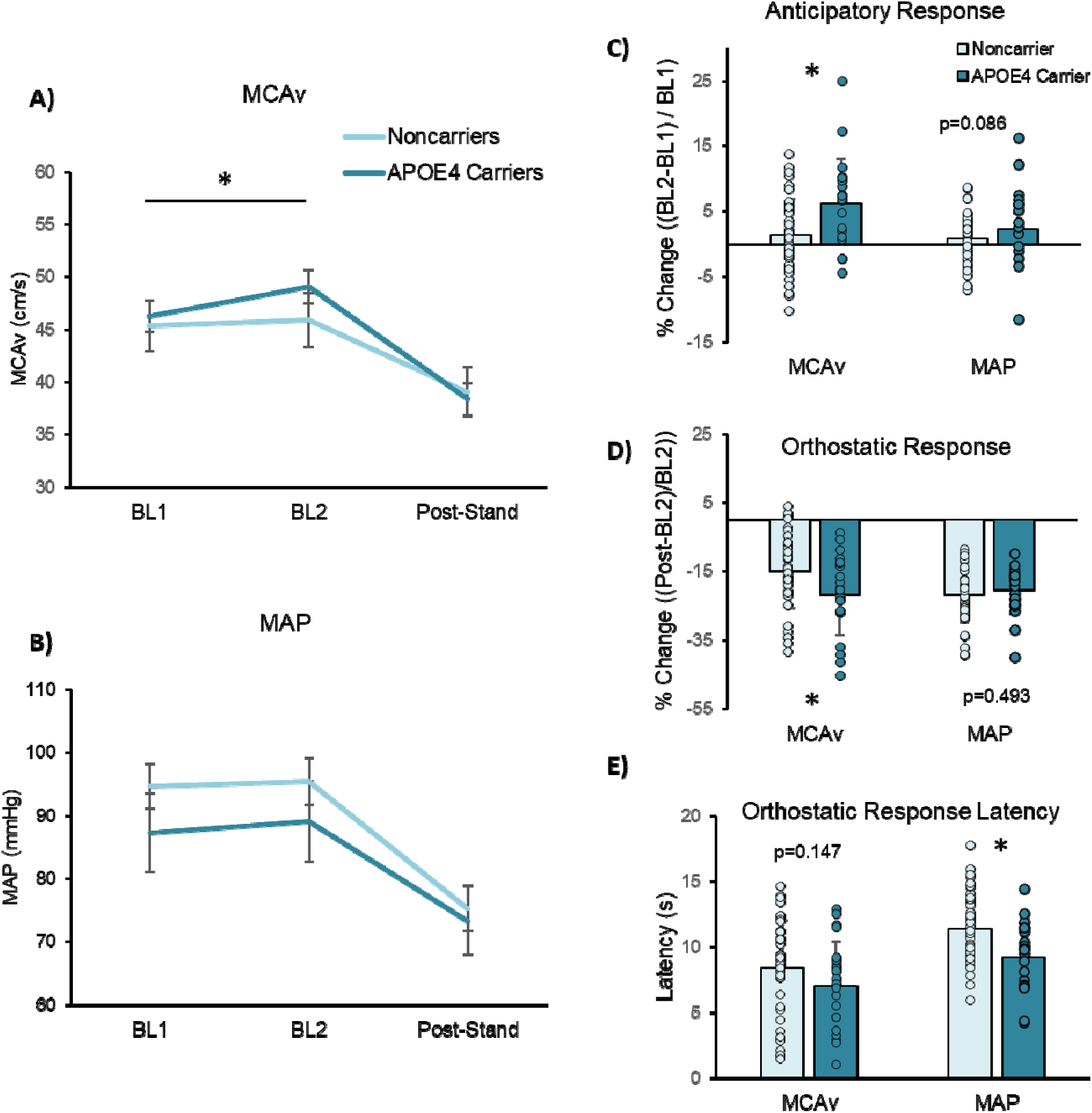
Cerebral and peripheral vascular responses during orthostasis in *APOE4* carriers and noncarriers with normal cognition. **(A)** Both genotypes experienced significant drops in middle cerebral artery blood flow velocity (MCAv) after standing. However, only *APOE4* carriers showed an anticipatory increase in MCAv just before standing, while noncarriers did not show any significant change. **(B)** Anticipatory increases in beat-to-beat mean arterial pressure (MAP) and drops in MAP after standing were observed across all participants, regardless of genotype **(C)** *APOE4* carriers demonstrated a greater anticipatory increase in MCAv but no difference MAP compared to noncarriers. **(D)** *APOE4* carriers showed greater post-stand decreases in MCAv compared to noncarriers, but no difference in MAP decreases. **(E)** *APOE4* carriers showed shorter MAP responses latencies (p=0.003) compared to noncarriers, while MCAv response latency was not different between genotypes (p=0.147) * indicates p<0.05 for between-group (genotype) differences.

For peripheral MAP, cognitively-normal *APOE4* carriers showed no difference in anticipatory change or orthostatic-induced decrease in MAP (**Figure 1B**), but did demonstrate shorter latencies of orthostatic-induced MAP responses (**Figure 1E**). There was no significant time-by-genotype interaction (F_2,124_ = 2.27, *p*=0.108) or main effect of genotype (F_2,124_ = 0.35, *p*=0.559). There was a main effect of time (F_2,124_ = 298.22, *p*<0.001), in which MAP was higher at BL2 compared to BL1 regardless of genotype (*p*=0.031), and was lower at the post-stand time point regardless of genotype (p<0.001). When comparing the normalized magnitude of change in MAP between groups, there were no significant differences in MAP anticipatory increase (*p*=0.086) (**Figure 1C**) or post-stand decrease (*p*=0.493) between groups (**Figure 1D**). *APOE4* carriers demonstrated shorter MAP response latencies compared to noncarriers (*APOE4* = 9.2 ± 2.7s; noncarriers = 11.4 ± 2.6s, *p*=0.003) (**Figure 1E**).

### 3.2. Effect of Alzheimer’s disease progression on cerebrovascular response to orthostasis in *APOE4* carriers and noncarriers

In *APOE4* carriers, Alzheimer’s disease progression (MCI and AD) was characterized by a loss of anticipatory increase in MCAv and slower MCAv responses to orthostasis compared to NC, while no effect of diagnosis was present in noncarriers. When testing whether MCAv differed over each time point among diagnosis groups, there were no significant interaction effects for either noncarriers (**Figure 2A**) or *APOE4* carriers (**Figure 2B**) (p>0.258), but there were significant main effects of time for both genotypes (p<0.001). Both genotypes and all diagnostic subgroups displayed a significant reduction in MCAv between BL2 and post-stand time points in response to orthostasis (p<0.001). While not statistically significant, noncarriers showed a trend for greater MCAv at BL2 compared to BL1 within the MCI subgroup (p=0.055), and a trend for MCI showing greater MCAv at BL1 compared to AD and NC (p≥0.065). NC *APOE4* carriers showed a significant MCAv increase between BL1 and BL2 (p<0.001), while MCI and AD A*POE4* carriers showed no statistical difference (MCI, p=0.079; AD, p=0.096).

**Figure 2.**
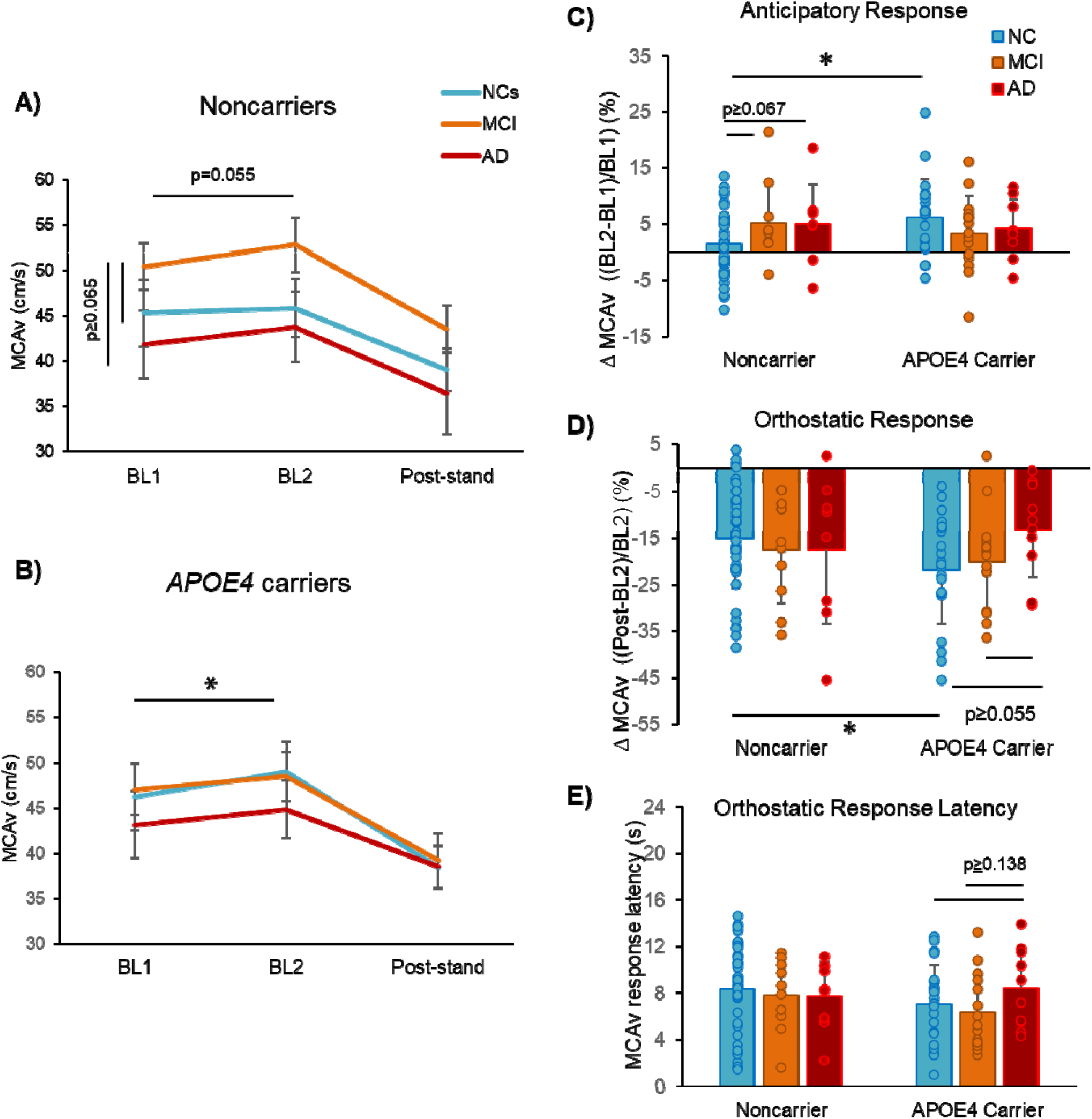
Cerebral vascular responses during orthostasis in over the course of AD progression in normal cognition (NC), mild cognitive impairment (MCI), and early-stage Alzheimer’s disease (AD). **(A)** Noncarriers of *APOE4* showed no significant effect of diagnosis on MCAv. **B)** NC *APOE4* carriers showed a significant anticipatory increase in MCAv, while this anticipatory response was attenuated in MCI and AD *APOE4* carriers. **(C)** NC *APOE4* carriers showed greater anticipatory percent increase in MCAv compared to noncarriers, but genotype differences were not observed in MCI and AD. **(D)** NC *APOE4* carriers showed greater orthostatic-induced reduction in MCAv, but no genotype differences were observed in MCI and AD. **(E)** No genotype or diagnoses differences were observed for MCAv response latency during orthostasis. *indicates p<0.05 for between-diagnosis and between-genotype differences.

For anticipatory %change in MCAv, there was a diagnosis-by-genotype interaction effect (F_5,105_ = 3.05, *p*=0.026) and no main effects of diagnosis or genotype (p>0.521). NC *APOE4* carriers showed greater anticipatory increases in MCAv compared to NC noncarriers (p=0.004), but no difference in anticipatory MCAv increases compared to noncarriers at the MCI and early AD stages of disease (p>0.490) (**Figure 2C**). In contrast, noncarriers with MCI showed a greater anticipatory MCAv increase on average compared to noncarrier NCs, though this difference did not meet our adopted level of significance (p=0.067).

For orthostatic-induced MCAv response, no diagnosis-by-time interaction or main effects were observed for the magnitude of MCAv change (**Figure 2D**) or latency of response (**Figure 2E**) (p>0.192). While NC *APOE4* carriers showed greater reduction in MCAv compared to noncarriers (**Figure 1D)** no group differences were observed at the MCI and early AD stages of disease (p>0.480) (**Figure 2D**). *APOE4* carriers with early-stage AD tended to show slower MCAv response latencies compared to MCI and NCs, but this difference did not meet our adopted level of significance (p≥0.138) (**Figure 2E**).

### 3.3. Effect of Alzheimer’s disease progression on peripheral blood pressure response to orthostasis in *APOE4* carriers and noncarriers

Disease progression did not affect the magnitude of anticipatory or orthostatic-induced changes in peripheral MAP in either *APOE4* carriers or noncarriers. When testing whether MAP differed over each time point between diagnosis groups, there were no significant interaction effects for either noncarriers (**Figure 3A**) or *APOE4* carriers (**Figure 3B**) (p>0.489), but main effects of time were present for both genotypes (p<0.001). Both genotypes and all diagnostic subgroups displayed a significant reduction in MAP between BL2 and post-stand time points in response to orthostasis (p<0.001). Noncarriers showed no difference at BL2 compared to BL1 within each diagnostic subgroup (p≥0.143). NC *APOE4* carriers showed a significant MAP increase between BL1 and BL2 (p<0.001), while MCI and AD A*POE4* carriers showed no statistical difference (MCI, p=0.136; AD, p=0.575).

**Figure 3.**
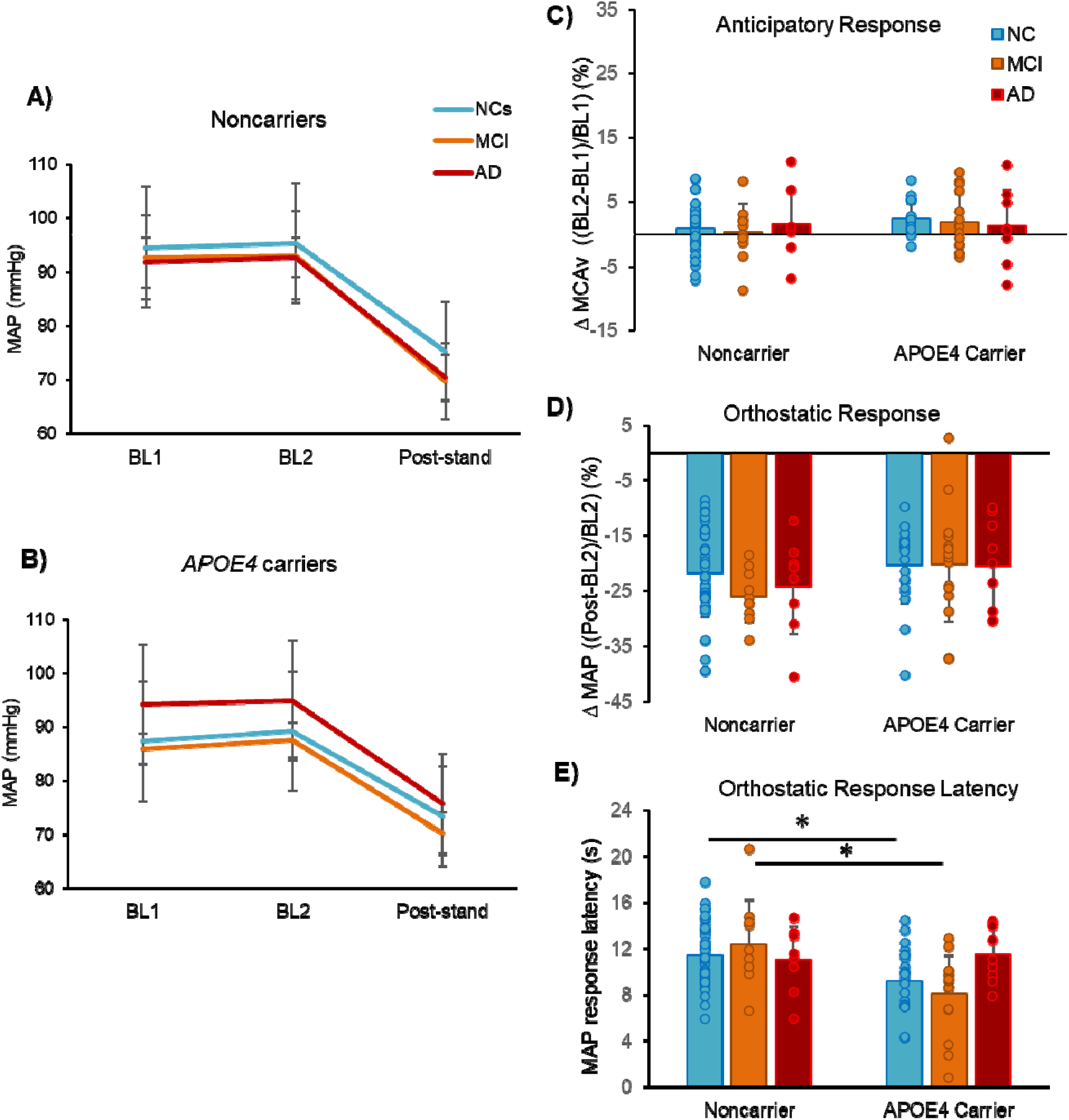
Mean arterial blood pressure (MAP) responses during orthostasis in over the course of AD progression in normal cognition (NC), mild cognitive impairment (MCI), and early-stage Alzheimer’s disease (AD). **(A)** In noncarriers, MAP decreased at post-stand time points in response to orthostasis but showed no difference in anticipatory increase, regardless of diagnosis. **B)** In APOE4 carriers, there was an anticipatory increase in MAP only in NCs. **(C)** Anticipatory MAP, nor **(D)** orthostatic-induced change in MAP differed by diagnosis. **(E)** Shorter latency of orthostatic-induced MAP response observed in NC *APOE4* carriers compared to noncarriers was also present at the MCI disease stage, but showed no difference in early AD. *indicates p<0.05 for between-diagnosis and between-genotype differences.

There were no diagnosis-by-genotype interaction effects for either anticipatory or orthostatic-induced change in MAP (p>0.707). There was a main effect of genotype, in which *APOE4* carriers showed greater anticipatory increases in MAP (p=0.040) and less reduction in MAP during orthostasis compared to noncarriers (p=0.004). There were no main effects of diagnosis for either anticipatory of orthostatic-induced changes in MAP (p>0.416).

There was a significant disease-by-genotype interaction for orthostatic MAP response latency (F_5,105_ = 3.42, *p*=0.037), in which the shorter latency of orthostatic-induced MAP response observed in NC *APOE4* carriers compared to noncarriers (**Figure 1E**) was also present at the MCI disease stage (*p*=0.007), but increased to comparable levels to noncarriers in early AD (*p*=0.743) (**Figure 3E**).

## 4. DISCUSSION

This study provides novel insights into the effects of the *APOE* genotype on mechanistic regulation of cerebral and peripheral vascular responses to orthostasis over the early stages of Alzheimer’s disease progression. The sit-to-stand paradigm provided a hemodynamic challenge to the cerebral vascular system[14–16,46] that exposed differences in cerebrovascular regulation between *APOE4* carriers and noncarriers. Here, cognitively-normal *APOE4* carriers showed greater drops in MCAv during orthostasis as well as higher anticipatory increases in MCAv and faster peripheral MAP responses compared to their noncarrier peers, which may reflect genotype-specific vascular adaptations to counteract an impaired cerebral pressure/flow relationship during orthostasis. Consistent with previous research in AD, we found that baseline resting cerebral blood velocity tended to show a (nonsignificant) decrease over disease progression.[29,47,48] However, the magnitude of orthostatic-induced drops in MCAv and MAP remained consistent [28] and even showed a trend towards attenuation (lesser drop) in MCI and AD diagnoses compared to NCs (**Figure 2D**, **Figure 3D**), potentially reflecting heightened sympathetic drive observed across the AD progression.[49–51] Together, our findings extend the knowledge of Alzheimer’s disease-related impairments and progression in cerebral vascular regulation to understand differential physiologic responses in individuals who carry the *APOE4* allele within hemodynamic behavioral contexts, which may play a role in their increased vulnerability to AD.[52] Importantly, our findings identify vascular compensatory strategies in *APOE4* carriers that may serve as a target for treatment efforts during this window of therapeutic opportunity in the prodromal stages of the disease.

The present results provide novel evidence that *APOE4* carriers utilize vascular compensation strategies that may counteract genotype-specific impairments in cerebral vascular regulation in the preclinical stages of AD. Greater anticipatory increases in MCAv (**Figure 1C**) in *APOE4* carriers may potentially serve as compensatory neurovascular adaptations to chronically impaired cerebral vascular regulation. This anticipatory increase in MCAv is consistent with greater recruitment of prefrontal cortical regions during mobility in cognitively-normal, older adult *APOE4* carriers,^10^ and heightened excitability of the prefrontal cortex in patients with AD.[53,54] Notably, the prefrontal cortex can strongly influence the regulation of cardiovascular function.^12,13^ Differences in anticipatory MCAv increase were not present in MCI and AD between genotypes (**Figure 2C**), implicating that these compensatory adaptations may be lost as *APOE4* carriers progress into clinical syndrome. Greater increases in anticipatory MCAv may reflect greater cerebral vascular contractility, resulting in exaggerated changes in vasoconstriction and dilation in response to changes in blood pressure, which has been reported in post-mortem examination of AD cortical tissue.[55] In the present study, greater cerebral vasomotor activity may be engaged in anticipation of hemodynamic blood flow reduction, resulting in more effective cerebral vascular regulation during orthostasis (**Figure 1A**). However, higher cerebral vasomotor activity could also lead to chronically reduced cerebral blood flow, especially if blood pressure becomes elevated.[55] Supporting this notion, there was no difference in baseline MCAv between cognitively-normal *APOE4* carriers and noncarriers (**Figure 1A**), but *APOE4* carriers with AD presented with higher baseline (BL1) MAP on average (**Figure 3B**). Greater anticipatory vascular compensation for hemodynamic dysfunction may also be consistent with recent research implicating that *APOE4* drives AD processes through a gain of abnormal neuronal function, rather than a loss of normal function.[56] However, without longitudinal assessments, it remains possible that older adults in the NC group reflect a “healthy survivor” bias, in which greater anticipatory MCAv and faster orthostatic MAP responses potentially contribute to increased neurocognitive resilience.[43] Future studies measuring cortical activity and that employ targeted modulation of key brain regions will help elucidate underlying neural mechanisms that may explain these differences in cerebral and peripheral anticipatory vascular responses in *APOE4* carriers.

Our findings provide evidence that impaired vascular responses to hemodynamic challenge in *APOE4* carriers are cerebral specific. We observed no differences in the magnitude of orthostatic-induced peripheral MAP changes between genotypes or across all stages of disease diagnosis (**Figure 3D**). Faster peripheral vascular responses to orthostasis in cognitively-normal *APOE4* carriers, indicated by shorter MAP response latencies (**Figure 1E**), may also be consistent with vascular compensation for greater orthostatic-induced drops in MCAv in this subgroup (**Figure 1D**). Similar to anticipatory cerebral vascular responses, we found that faster MAP responses during orthostasis were present only in cognitively-normal older adults and slowed in older adults with AD (**Figure 3E**). These results could be explained by a heightened sensitivity of the arterial baroreflex in cognitively-normal *APOE4* carriers that becomes gradually desensitized over time.[57] Autonomic dysfunction of blood pressure regulation has been associated with AD pathology to the insular cortex, which may negatively affect baroreflex mechanisms of blood pressure control.[58] The present results support distinct phenotypes of cerebral vascular dysfunction in *APOE4* carriers and noncarriers throughout the course of AD progression, in which peripheral vascular responses may act synergistically with greater anticipatory cerebral vascular mechanisms as neuroprotective features in the prodromal disease stage.

We observed two unexpected findings involving noncarriers in this study: 1) anticipatory MCAv tended to increase in the MCI stage of disease (**Figure 1C**) (though did not reach statistical significance p=0.055) and 2) baseline (BL1) MCAv in noncarriers with MCI tended to be higher than NCs and AD (p≥0.065) (**Figure 1A**). While these patterns did not reach statistical significance in the present study, they may identify directions for future investigation involving dissociable effects of *APOE4* in the MCI stage of AD processes. While a decrease in cerebral blood flow is a consistent finding in AD, [29,47,48] a paradoxical increase in cerebral perfusion has also been reported in the early stages of neurodegenerative diseases such as Parkinson’s disease.[59] This initial period of increased cerebral perfusion is posited to be a compensatory response for the emergence of orthostatic hypotension in these patient populations.[59] Consistent with this hypothesis, noncarriers with MCI and AD in the present study tended to show greater orthostatic-induced drops in MCAv (**Figure 2C**) and MAP (**Figure 3C**) compared to noncarrier NCs, which could have influenced their tendency for higher levels of cerebral blood velocityobserved at baseline (**Figure 2A**). Our results therefore suggest that *APOE4* carriers and noncarriers may present two distinct phenotypes of cerebral vascular dysfunction in AD progression, in which *APOE4* carriers display a loss of effective vascular compensation, while noncarriers display an engagement in compensation during early stages of the disease (MCI) that may be less effective in resisting clinical syndrome. Future studies are needed to determine the effectiveness of targeted treatments for brain vascular health in resisting cognitive decline in each *APOE* phenotype of cerebral vascular dysfunction.

### 4.1. Limitations

The biological variable of sex can interact with aging and brain vascular function to influence cognitive function;[12] thus, differences in sexes in the NC group in the present study should be considered in the interpretation of the present results. While previous studies using MR-based imaging show no changes in cerebral vessel diameter in response to change in P_ET_CO_2_,[60] it is possible that changes in MCA vessel diameter could have influenced the present results and were not captured in our TCD measures of MCAv. People with MCI and AD may be increasingly prescribed antihypertensive medications,[28,61] which may have an effect on cerebral and peripheral responses to orthostasis and could not be controlled for in the present study. During the sit-to-stand, other factors including neurovascular coupling, sympathetic activity, and cardiac output can affect cerebral and peripheral responses to orthostasis and were not captured in the present study.

### 4.2. Conclusions

For the first time, our findings show that hemodynamic challenge exposes *APOE* genotype-specific deficits in cerebral vascular responses to orthostasis in older adults who carry the E4-allele. Our findings also reveal greater anticipatory increases in cerebral blood velocity and faster arterial pressure responses to orthostasis, consistent with vascular compensatory mechanisms, in cognitively-normal *APOE4* carriers that may be lost as AD progresses. Further, differences in the trajectories of cerebral and peripheral vascular function over the course of AD progression implicate that *APOE4* carriers and noncarriers present with different phenotypes of brain vascular function during hemodynamic challenge that may be clinically-relevant to cognitive decline. These findings may identify specific features of cerebral vascular dysfunction that could be targeted through precision-based approaches in individuals at high genetic risk for AD.

## Data Availability

All data produced in the present study are available upon reasonable request to the authors.

## ACKNOWLEDGMENTS

a. We wish to acknowledge Emily Hazen for assistance with data collection and analysis.
b. Funding: This work was supported by the National Institute on Aging of the National Institutes of Health [K99AG075255 (JP)] and [P30 AG072973 (SB)], the National Heart, Lung and Blood Institute [T32HL134643 (AWH)] the Cardiovascular Center’s A.O. Smith Fellowship Scholars Program, and the Georgia Holland Endowment Fund. The content of this publication is solely the responsibility of the authors and does not necessarily represent the official views of the National Institutes of Health or any other funding agency. The funding sources had no involvement in the collection, analysis, or interpretation of the presented data.

## CONSENT STATEMENT

All human subjects in this study provided written, informed consent.

